# ASSOCIATION BETWEEN CLIMATIC VARIABLES AND CARDIOVASCULAR HOSPITALIZATIONS IN BRAZIL: AN ECOLOGICAL STUDY

**DOI:** 10.1101/2025.09.23.25336510

**Authors:** Bruna Dóris, Gabriel Savogin Andraus, Rajesh da Silva Seunaraine, Débora Nabor de Cássia Silva, Natália da Silva Teixeira, Bruno Siegel Guerra, Thyago Proença de Moraes, Gustavo Lenci Marques

**Affiliations:** Universidade Federal do Paraná, Postgraduate Program in Internal Medicine and Health Sciences, Curitiba, Paraná - Brazil; Pontifícia Universidade Católica do Paraná, Curitiba, Paraná - Brazil; MUNAI, Curitiba, Paraná - Brazil

**Author notes:** Corresponding author: Bruna Dóris; 55 41 99640-2949.

**Keywords:** Brazil, cardiovascular diseases, hospitalization, incidence, temperature

## Abstract

Cardiovascular disease is the leading cause of death worldwide, and external factors such as ambient temperature can predispose to cardiovascular events. Brazil, a country with continental dimensions and diverse climates, lacks a recent study comparing cardiovascular hospitalization incidence with climatic data across the Brazilian states. This study aimed to describe and relate the incidence of cardiovascular events, including acute myocardial infarction, stroke, and heart failure, to climatic data. Data were obtained from the Department of Informatics of the Unified Health System and the National Meteorology Institute from 2010 to 2024. We recorded a total of 2,917,900 hospitalizations for cardiovascular diseases across 103 cities in Brazil where complete climatic data were available. The incidence per 100,000 inhabitants ranged from 0.007 in the state of Maranhão to 0.023 in Rio Grande do Sul. We observed a lower relative risk of hospitalization for cardiovascular diseases in regions with a higher ambient temperature, with the temperature range up to 19°C having the highest risk. No difference in hospitalization risk was observed between the temperate and tropical climate regions. This study highlights the importance of public measures for cardiovascular event prevention during periods of lower temperature throughout the year.

## INTRODUCTION

Cardiovascular disease is the leading cause of death in Brazil and worldwide, and its prevalence has increased in recent decades due to population aging.(de Oliveira et al., 2022, 2024; Institute for Health Metrics and Evaluation Population Health, 2022) External factors, such as pollution levels, stress, and climate, can influence the cardiovascular system. The latter has been widely studied and classically described in literature, with cardiovascular mortality rates expected to vary according to seasonal and daily temperature fluctuations. However, published studies on this topic have yielded divergent results, with some indicating a higher risk associated with heatwaves(Sen et al., 2016; Sun et al., 2018; Zhao et al., 2019) and others suggesting a higher risk with exposure to lower temperatures.(Barnett et al., 2005; Claeys et al., 2017; Ferreira et al., 2019; Gasparrini et al., 2015; Gerber et al., 2006; W. R. Keatinge, 1997).

The evidence supporting the risk of cardiovascular events with lower temperatures is more robust. It is well established that exposure to lower temperatures induces sympathetic hyperactivity, thereby increasing the cardiac workload. Furthermore, increased platelet activation and hemoconcentration have been observed, leading to a heightened risk of atherothrombosis(Claeys et al., 2017; W. R. Keatinge, 1997; William R Keatinge et al., 1986; Paulo et al., 2004). Research has shown that these effects can persist for up to four weeks following exposure to cold temperatures(Claeys et al., 2017; Gasparrini et al., 2015).

In this context, a German ecological study compared daily acute myocardial infarction (AMI) rates from 1987 to 2014 with average ambient temperatures. The results revealed that temperatures below 18·4 °C were associated with a higher relative risk of AMI(K. Chen et al., 2019). An American study analyzing data from 1979 to 2002 in Minnesota found seasonal variations in sudden cardiac death, with a peak in winter and a nadir in summer(Gerber et al., 2006). Another retrospective study demonstrated that winter was associated not only with higher infarction rates, but also with more extensive and severe events (Kloner et al., n.d.). Japanese researchers found a correlation between lower temperatures and an increased risk of hospitalization for cardiovascular reasons, particularly in the older adults (Rui Pan et al., 2023).Some studies suggest that the relationship between cardiovascular mortality and ambient temperature follows a U-shaped curve, with temperatures between 15 °C and 25 °C associated with the lowest mortality rates(Abrignani et al., 2022; Ferreira et al., 2019; Gasparrini et al., 2015; Liu et al., 2015).

However, establishing this correlation may be challenging in countries with continental proportions, such as Brazil. A 2019 Brazilian study evaluated the daily temperatures and myocardial infarction mortality in six capitals in different regions, climates, and socioeconomic conditions between 1996 and 2013. The results were consistent with those of international studies, which showed higher AMI rates in winter in locations with greater temperature amplitudes. However, in cities closer to the equator, myocardial infarction mortality did not exhibit a significant relationship with temperature (Ferreira et al., 2019). Another national study involving capitals from distinct regions found that the risk of hospital admission for cardiovascular reasons was lower during summer(Zhao et al., 2019). While these studies had significant impacts, they had limitations, such as focusing solely on myocardial infarction mortality in the capital or evaluating only periods of higher temperatures.

This study aimed to describe the incidence of cardiovascular events in Brazil from 2010 to 2024 and its association with daily average temperatures. Additionally, it investigated differences in the incidence of cardiovascular events between the temperate and tropical climate regions in the country.

## MATERIALS AND METHODS

This ecological study was based on the analysis of extensive national records with open data for public consultation. According to the Conselho Nacional de Saúde Resolution 510/2016, because this research used publicly accessible information databases in accordance with Law No. 12,527 of November 18, 2011, whose aggregated information does not allow individual identification, approval by the Research Ethics Committee/National Research Ethics Committee was not required.

### Inclusion criteria

Data from the Department of Informatics of the Unified Health System (DATASUS) were directly extracted from the national system using the TabWin® system to obtain epidemiological data. For this analysis, all hospitalization rates by residential municipality from January 2010 to June 2024 were included for the following International Classification of Diseases (ICD)-10 codes: acute myocardial infarction (ICD-10: I21, I22), other ischemic heart diseases (ICD-10: I20, I23-I25), heart conduction disorders and cardiac arrhythmias (ICD-10: I44-I49), heart failure (ICD-10: I50), cerebral infarction (ICD-10: I63), and stroke (ICD-10: I64).

Meteorological data were automatically extracted from the National Meteorological Institute. Data from all Brazilian meteorological stations, including hourly average temperatures, were obtained from January 2010 to June 2024. All extracted data were converted from their native format to the comma-separated value (CSV) format, generating an. csv file for better interpretation and analysis using R.

### Exclusion criteria

Cities with a lower number of events than the number of observation days (5296) were excluded to ensure the convergence and stability of the statistical models. Furthermore, cities with intermittent or sparse data were removed to avoid compromising the validity of the estimated lag effects and seasonal trends.

### Statistical analysis

Initially, publicly available data from the Meteorological System and DATASUS were processed, cleaned, and categorized. The variables of interest for the analysis were the daily hospitalization counts (Count) and the daily mean temperature (DAILY_MEAN_TEMP) for each municipality (MUNIC_MOV). Daily mean temperature data (DAILY_MEAN_TEMP) were obtained from [Number or Source, e.g., a network of surface weather stations] for each city and processed to generate a single representative time series for the urban area. For correct geographical allocation and regional grouping, the data were merged with spatial information from the geobr package, which allowed for the disambiguation of capital cities with identical names (e.g., Belém-PA and Belém-AL). To ensure the integrity of the modeling, only cities that presented continuous time series, without gaps in the daily records of hospitalizations and mean temperature during the study period, were included in the analysis.

The distribution of quantitative variables was checked using histograms, and results are reported as mean (± standard deviation) for normally distributed data or median (minimum – maximum) for the non-normally distributed data. For qualitative variables, values for each group are expressed as absolute numbers (n) and percentages (%).

To model the association between daily mean temperature and the incidence of cardiovascular hospitalizations across different Brazilian cities, a two-stage data analysis approach was applied, as recommended for multi-city studies (Alahmad et al., 2023). In the first stage, the association was assessed for each city individually; in the second stage, the city-specific results were combined through a multivariate meta-analysis to generate pooled estimates for the macro-regions and for Brazil as a whole.

### First stage

A generalized linear model (GLM) with a quasi-Poisson family was fitted to the daily hospitalization counts. This approach was chosen to account for overdispersion commonly found in health count data. To model the non-linear and delayed effects of temperature, a distributed lag non-linear model (DLNM) was employed using the dlnm package for R. Details of the model are explained in the Appendices.

As a result of the first-stage analysis, the vector of coefficients (betas) and the variance-covariance matrix of the cumulative association were extracted for each city. From the estimated effect curve, the minimum morbidity temperature and the temperature of maximum effect were identified.

### Second stage

A multivariate meta-analysis was performed using the mvmeta package in R. The coefficients and covariance matrices from all cities were pooled to estimate an overall exposure-response curve. The selected estimation method was restricted maximum likelihood (REML). This process was repeated to pool the results by macro-region (North, Northeast, Central-West, Southeast, and South) and for Brazil. In addition, a meta-regression was conducted to evaluate the effect of climate type, by categorizing cities as “Temperate” or “Tropical” and using this variable as a moderator in the meta-analysis model. Notably, the city of Belém was specifically excluded from the pooled analysis of the North Region, as detailed in the analysis script.

All statistical analyses, as well as the creation of graphs and tables, were performed using R software (version 4.0.0 or higher), with intensive use of the dlnm, mvmeta, splines, mgcv, and tidyverse packages.

## RESULTS

Between January 2010 and June 2024, 2,917,900 hospitalizations for cardiovascular diseases were registered in Brazil in 103 cities with fully available climate data. The total number of hospitalizations in Brazil was significantly higher (approximately 20,000,000 cardiovascular-related hospitalizations). However, these data were excluded because not all Brazilian cities had complete climatic data (Figure 1). Additionally, cities with fewer cardiovascular hospitalizations during the analysis period than on the days of observation (5296 days) were also excluded. All state capitals were included in this study.

**Figure 1.**
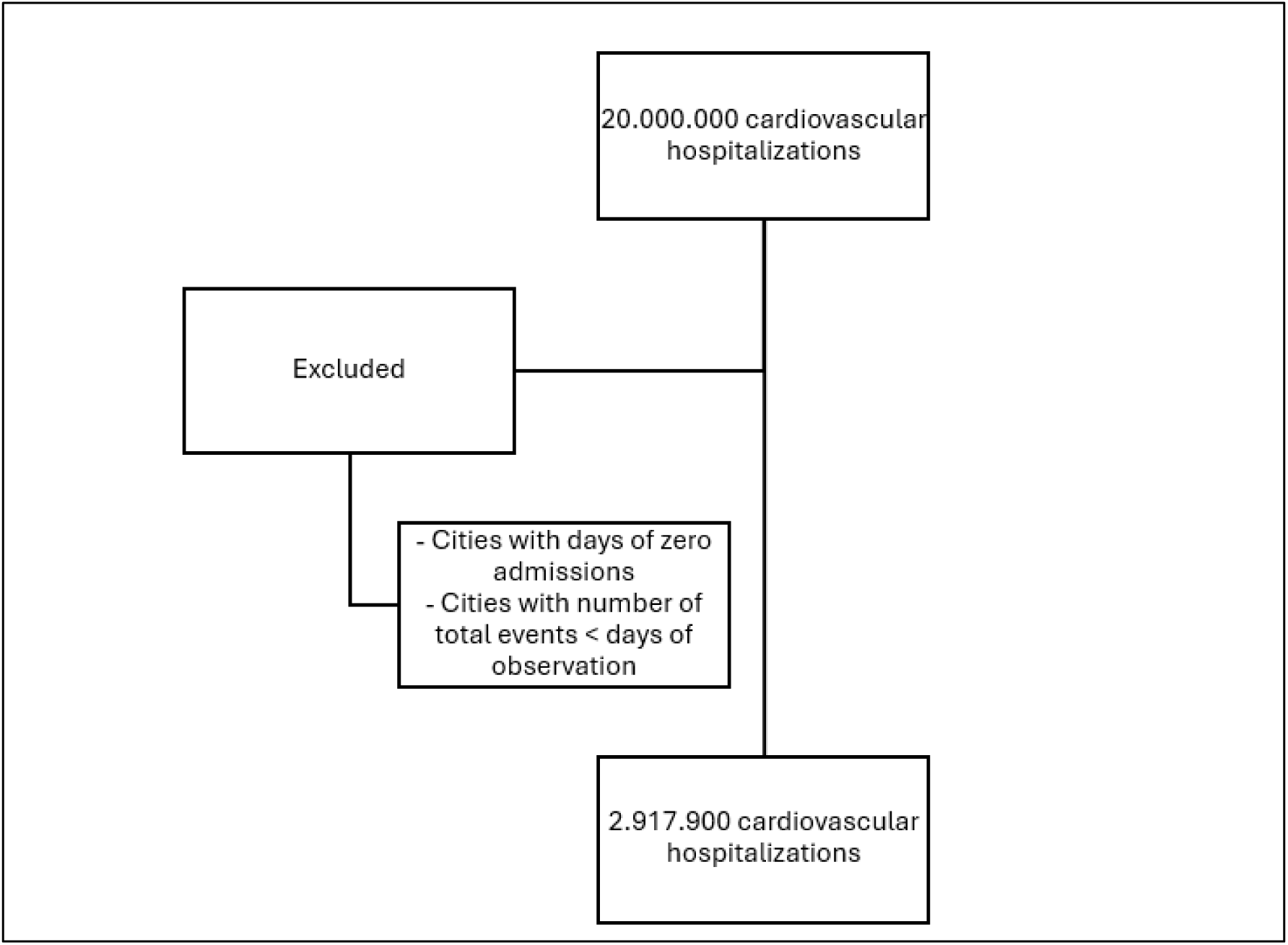
Flowchart of the Department of Informatics of the Unified Health System (DATASUS) data collection.

We calculated the incidence of cardiovascular diseases in each state during this period based on the number of hospitalizations and number of inhabitants in the state according to the current Instituto Brasileiro de Geografia e Estatística registration for 2024. The lowest incidence was in the northeastern region (0.007 in the state of Maranhão), whereas the highest was in the southern region (0.023 in the state of Rio Grande do Sul). The complete incidence list by state is provided in the Supplementary Material.

We mapped the incidence of hospitalization due to cardiovascular diseases per 100,000 inhabitants in each Brazilian state, as illustrated in Figure 2.

**Figure 2.**
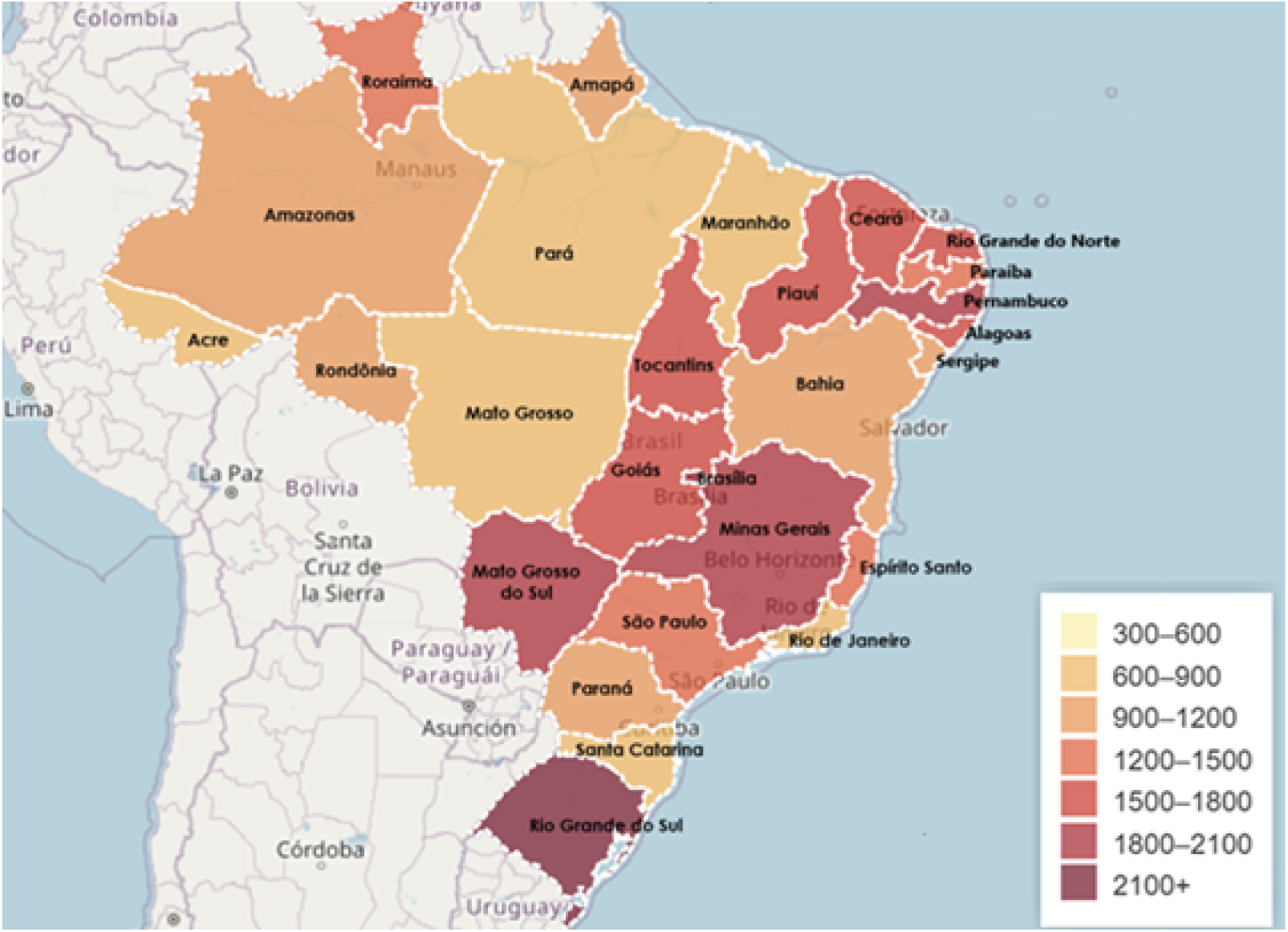
Incidence of cardiovascular hospitalizations per 100,000 residents from 2010-2024 for each state in Brazil.

We plotted the data comparing the daily median temperature and the relative risk of cardiovascular hospitalization (Figure 3).

**Figure 3.**
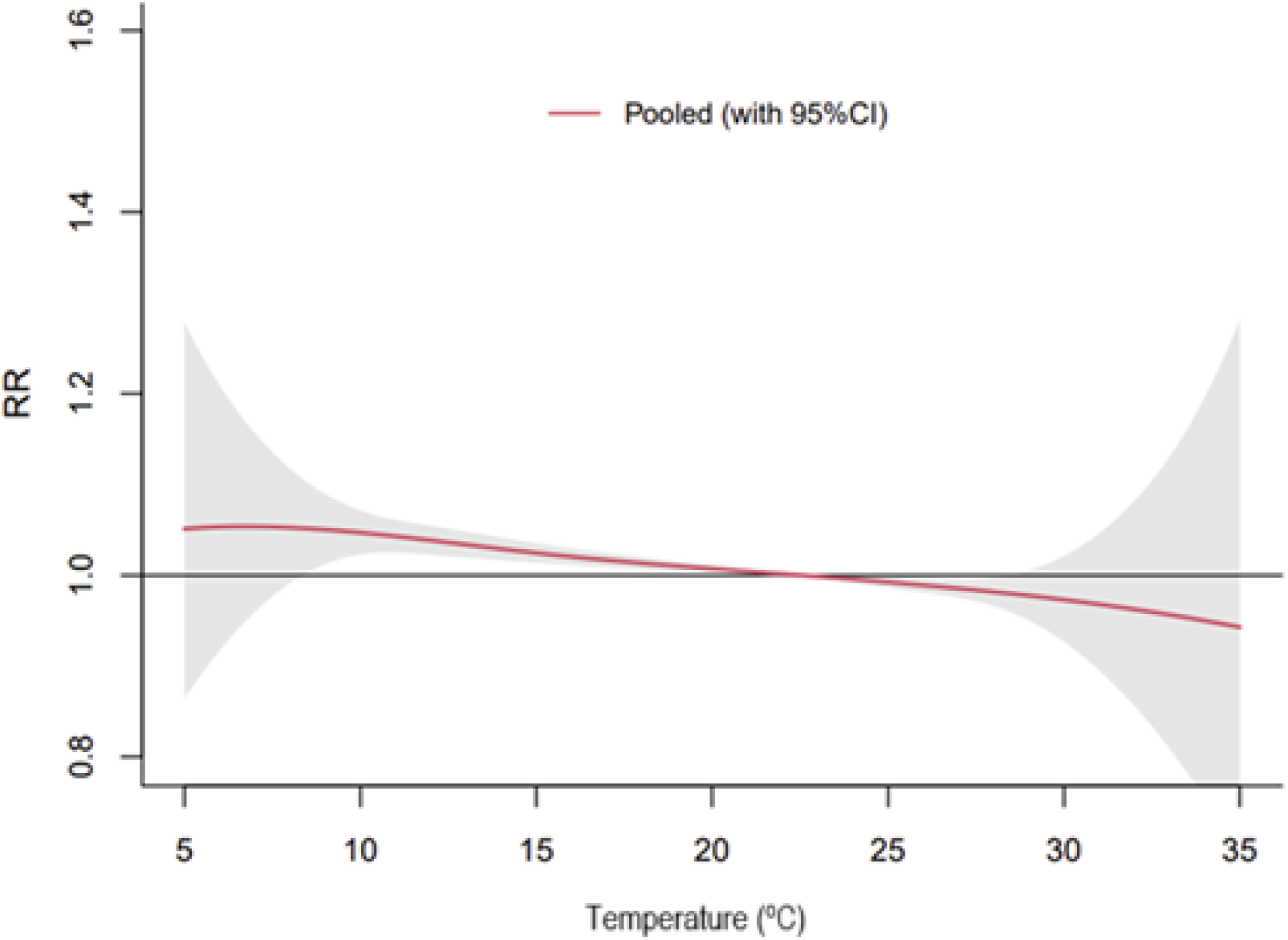
Relative risk of cardiovascular hospitalization according to daily median temperature in Brazil from 2010 to 2024. RR, relative risk; CI, confidence interval.

In the lowest temperature range (between 5 °C and 8 °C), there was a trend towards a higher incidence of hospitalizations. Because few Brazilian cities reach lower temperatures, the confidence interval of the relative risk was broad and included 1.0. In this zone, the relative risk of hospitalization was 1.05, with the 95% confidence interval decreasing as the temperature increased: [0.86– 1.28] at 5°C and [0.99–1.12] at 8°C.

Between temperatures of 9°C to 18°C, we observed a higher relative risk of hospital admission, with the risk increasing as the temperature decreased: at 9°C, the relative risk was 1.05 [1.01–1.09], at 14°C it was 1.03 [1.02–1.04], and at 18°C it was 1.01 [1.01–1.02].

From 19°C to 24°C, we observed a neutral relationship regarding hospitalization incidence and ambient temperature. Some authors advocate an ideal temperature range for the cardiovascular system, and according to our study, temperatures above this range may be considered the ideal range in Brazil. Above 25°C, we observed a trend of a protective effect from heat concerning cardiovascular hospitalizations. As the temperature increased, the relative risk of hospitalization decreased: the risk was 0.99 [0.99–1.0] at 25°C, 0.97 [0.93–1.02] at 30°C, and 0.94 [0.69–1.28] at 35°C. In parallel with the lower temperature extremes, since few cities had sufficient data for daily medium temperatures above 25°C, the confidence interval crossed 1.0.

As Brazil is a country of continental dimensions and most studies on the subject involve temperate climate countries, we compared the regions of temperate climate with those with tropical climate (Figure 4).

**Figure 4.**
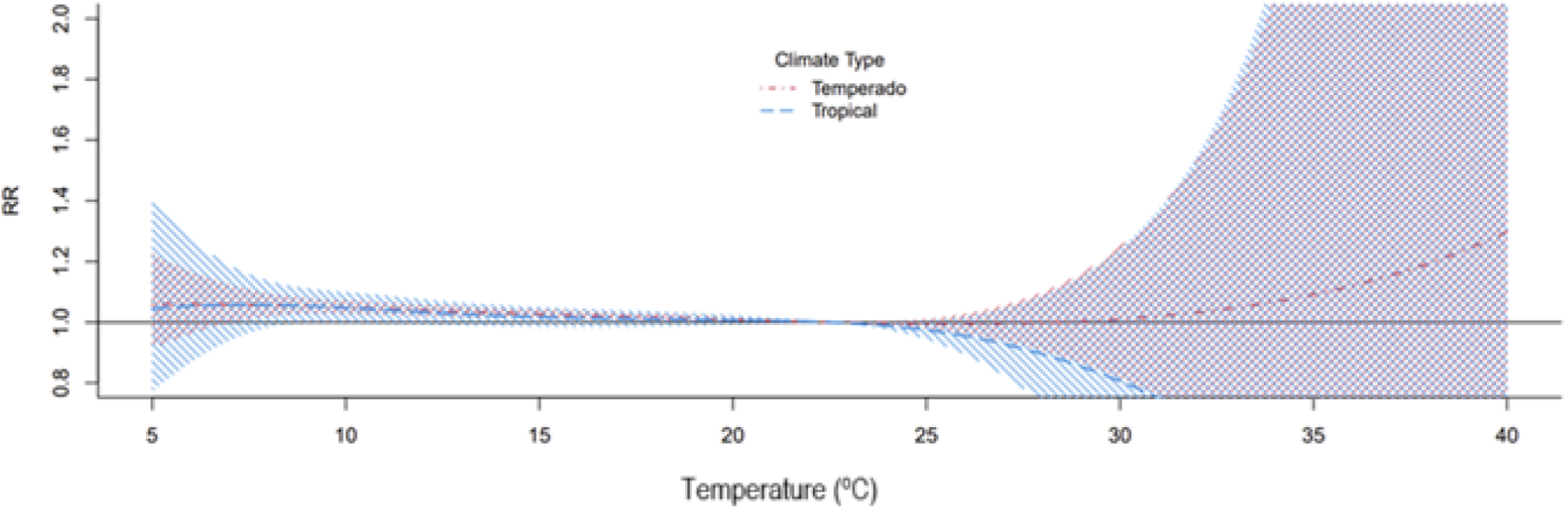
Relative risk of cardiovascular hospitalization according to the type of climate in Brazil during the period of 2010–2024. Shading indicates 95% confidence interval (CI). RR, relative risk.

No significant difference in hospitalization rates for cardiovascular diseases was observed between the temperate and tropical climate regions. We noted a tendency for a higher hospitalization risk in both climates at lower temperatures. Between 20°C and 24°C, ambient temperature had a neutral effect on hospitalization incidence. Above 25°C, a trend toward a lower risk of hospitalization was observed. The confidence intervals increased with increasing temperature, probably because of the smaller amount of data at extreme temperatures.

## DISCUSSION

To the best of our knowledge, this is the largest study conducted in Brazil that associates hospitalization for cardiovascular diseases with mean daily ambient temperature. Previous national studies have related climate data to cardiovascular mortality or have limitations, such as focusing on a specific cardiovascular disease, restricted time periods, or only covering capital cities.

The relationship between cardiovascular events and ambient temperature has been firmly established at lower temperatures, and our findings align with those reported in the literature. Decreased body temperature leads to vasoconstriction, increased blood pressure, heightened skeletal muscle tone for heat conservation, and an increased concentration of vasoconstrictor peptides. Consequently, the cardiac workload increases. Moreover, there may be greater crystallization of atherosclerotic plaques, which increases the risk of plaque rupture (de Oliveira et al., 2022, 2024; Institute for Health Metrics and Evaluation Population Health, 2022; Sen et al., 2016; Sun et al., 2018; Zhao et al., 2019).

Our study encompassed various cardiovascular diseases and highlighted the association between hospitalization due to cardiovascular diseases and temperatures below 19°C. This finding is similar to that reported by Chen et al. in Germany, where the risk of hospitalization due to heart attack was higher until a temperature of 18·4°C (W. R. Keatinge, 1997). Katayama et al. assessed the tomographic characteristics of coronary plaques responsible for heart attacks in 202 patients in Japan and found a higher incidence of plaque rupture at lower temperatures(Katayama et al., 2020). In that study, the average temperature at the peak of heart attacks was 16.5°C, with a higher risk of myocardial infarction between 10°C and 20°C.

Although the association between ambient temperature and other cardiovascular diseases in the literature is noteworthy, it is less pronounced than that with coronary artery disease(Cheng et al., 2010; Khraishah et al., 2022). An ecological study conducted in Taiwan analyzed data from 2012 to 2019 and found higher rates of first decompensation for heart failure during the winter months, especially in patients aged > 60 years with at least one comorbidity(D.-Y. Chen et al., 2024). Alahmad et al. conducted a multinational study exploring the association between cardiovascular event mortality in individuals over 40 years of age and ambient temperature in 27 countries. They found higher mortality due to heart failure at extreme temperatures(Alahmad et al., 2023). Hospitalization rates for heart failure decompensation are higher in winter; however, this also occurs alongside higher rates of respiratory infections.(Alahmad et al., 2023; Stewart et al., 2002) A multinational ecological study estimated that for every 1,000 deaths from stroke, nine were attributed to extreme cold and 1.6 to extreme heat (Alahmad et al., 2023). Studies on stroke etiology are inconsistent, but generally, there is a greater risk of events of both etiologies (ischemic and hemorrhagic) at temperature extremes(J. hua Chen et al., 2017; Cheng et al., 2010; Feigin et al., 2021; Lavados et al., 2018).(J. hua Chen et al., 2017; Cheng et al., 2010; Feigin et al., 2021; Lavados et al., 2018) As cardiac arrhythmia is a broad term that encompasses various pathologies, studies on this topic are divergent (Alahmad et al., 2023; Khraishah et al., 2022).(Alahmad et al., 2023; Khraishah et al., 2022) Alahmad et al. found an association between mortality from arrhythmias and lower temperatures; however, they did not find an association with higher temperatures (Alahmad et al., 2023). This may be attributed to misclassification since a ventricular arrhythmia may result from cardiac ischemia(Alahmad et al., 2023; Pimentel et al., n.d.). (Alahmad et al., 2023; Pimentel et al., n.d.)Some studies have indicated an association between atrial fibrillation and lower temperatures; however, the evidence is weak.(Nguyen et al., 2015; Zhu X et al., 2023)

We believe that in our study, hospitalizations for cardiovascular diseases as a whole were driven by coronary events. As our objective was to assess cardiovascular diseases, individual analyses of each ICD code were not conducted.

The number of coronary events over a 28-year period in Germany has been assessed, with stratification into two periods: 1987-2000 and 2001-2014. Similar to our study, cold was associated with higher rates of heart attack in both periods. Interestingly, in the more recent period, there was a greater influence of heat on these hospitalizations, and a warmer climate could be considered a potentially preventable trigger of heart attacks.(K. Chen et al., 2019)

Increased body temperature leads to dehydration and volume depletion, activating the sympathetic system, causing tachycardia and subsequently increasing the cardiac workload. Hemoconcentration also contributes to hypercoagulability, which increases the risk of thrombosis and heart attacks(Abrignani et al., 2022; Khraishah et al., 2022)(de Oliveira et al., 2022, 2024; Institute for Health Metrics and Evaluation Population Health, 2022).

Hundessa et al. found that of the total cardiovascular deaths worldwide from 2000 to 2019, 8.86% were caused by a non-optimal ambient temperature (high or low). However, only 0.66% were related to excessive heat. With climate change, it is suggested that warmer climates interfere with cardiovascular events(K. Chen et al., 2019; Hundessa et al., 2024; Khraishah et al., 2022; Rui Pan et al., 2023). However, we did not observe this effect in our study. The number of hospitalizations for cardiovascular diseases in Brazil reduced as ambient temperature increased, with heat potentially having a protective effect when temperatures exceeded 20 °C.

There were no statistically significant differences between the data for tropical and temperate cities. However, temperate climate regions showed a tendency toward a “U-shaped” curve, similar to that observed in the German study after the year 2000 and other reports on cardiovascular mortality.(Abrignani et al., 2022; Al-Kindi et al., 2023; Ferreira et al., 2019; Gasparrini et al., 2015; Khraishah et al., 2022; Paulo et al., 2004)

Our study is the first to compare regions with tropical climates with those with temperate climates in the same country. As shown in Figure 5, equatorial climate regions (a subtype of tropical climates) have the lowest incidence of hospitalization for cardiovascular diseases. We can formulate a hypothesis regarding the humidity of the Amazon rainforest. However, humidity data were not assessed in this study. In the South, which is the only temperate region of the country, the state with the highest incidence of cardiovascular hospitalization was Rio Grande do Sul. The eating habits in this region involve a diet high in fat; however, because our study did not evaluate this type of exposure, we cannot attribute it as a cause of more cardiovascular events.

**Figure 5.**
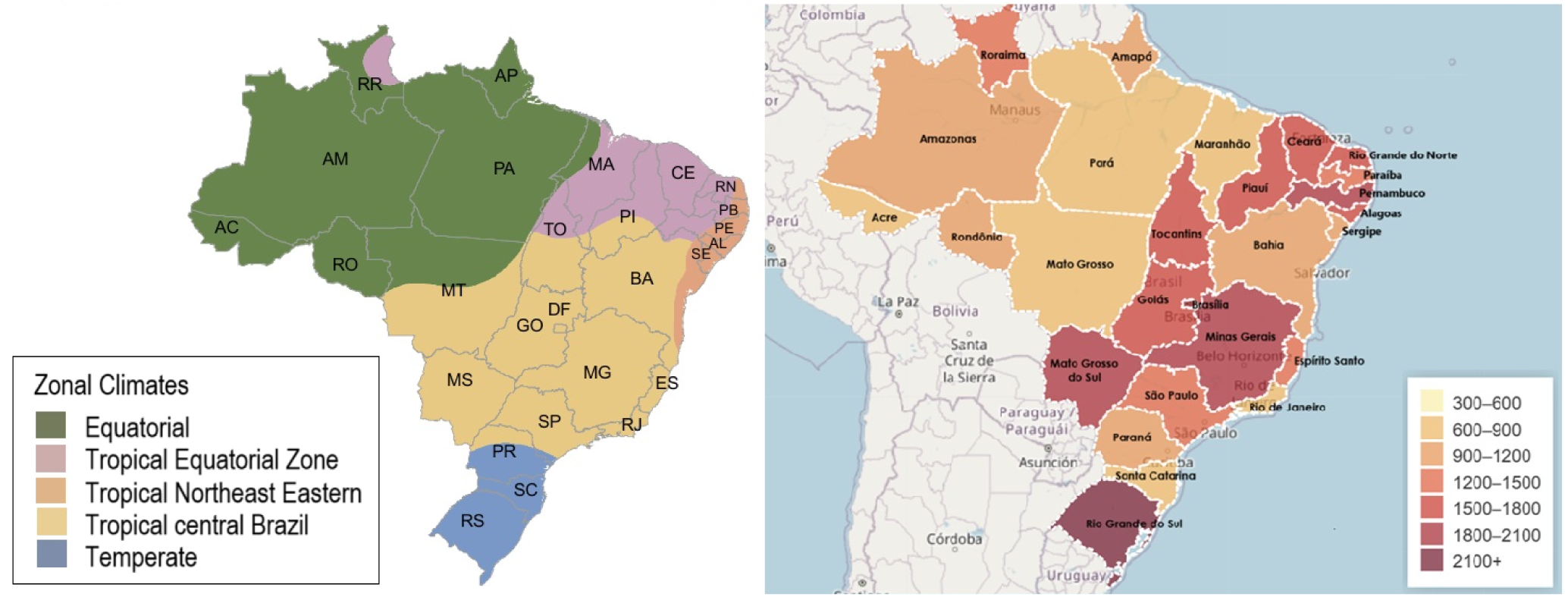
Comparison between climate maps* and incidence of hospitalization due to cardiovascular diseases per 100,000 inhabitants in Brazil. *Adapted from Instituto Brasileiro de Geografia e Estatística

The effects of temperature on the cardiovascular system vary according to the socioeconomic conditions. Populations with poorer socioeconomic conditions are more exposed to the effects of ambient temperature than are populations with better conditions.(Al-Kindi et al., 2023; Khraishah et al., 2022; Liu et al., 2015) This underscores the importance of public health measures to reduce the risk of cardiovascular hospitalization.

The effect of temperature on the cardiovascular system does not necessarily imply immediate repercussions. There can be a time lag in exposure to a certain temperature that culminates in a cardiovascular event. It is estimated that exposure to heat can have consequences on the cardiovascular system within 3 days, whereas exposure to cold can exert an effect on the cardiovascular system in a more delayed manner, from 7 to 21 days.(Abrignani et al., 2022; Gasparrini et al., 2015; Liu et al., 2015) In our study, we calculated a 7-day exposure period (high and low); however, the 7-day lag did not significantly influence hospitalization rates.

Under the current climate change scenario, this topic has gained importance as a public health measure for reducing cardiovascular morbidity and mortality. Public financial incentives for better home infrastructure, awareness campaigns, and increased financial support for health institutions focusing on cardiovascular treatment during extreme temperatures are examples of governmental measures that can be taken to mitigate climate consequences.(Feigin et al., 2021; Khraishah et al., 2022)

This study has some limitations. As this was an ecological study, we cannot assert a causal relationship between hospitalization for cardiovascular diseases and lower temperatures. Various other well-established factors can affect the cardiovascular system, such as humidity, pollution, and diet,(Murray et al., 2020) which have not been considered. Hospitalization data were collected from a national database, with data added at the time of each patient’s hospitalization. Thus, the ICD codes may have been incorrectly inputted by the attending physician. The initial objective of this research was to evaluate all cities in Brazil; however, climate data for smaller cities were incomplete or the number of hospitalizations for the evaluated ICD codes was lower than the number of days assessed. Consequently, most Brazilian cities were excluded from the analysis.

A positive aspect of our study is that it is the first Brazilian study to associate ambient temperature with hospitalization for cardiovascular diseases across more than 100 cities throughout the national territory, using updated data from the last 14 years. Furthermore, this is the only national study to compare data from cities with temperate and tropical climates.

In conclusion, from January 2010 to June 2024, ambient temperatures below 19°C in Brazil were associated with increased hospitalizations due to cardiovascular diseases. No significant differences were observed between the tropical and temperate cities. This association may be altered by climate change, emphasizing the need for public health measures to reduce the risk of cardiovascular hospitalization.

## Data Availability

Raw data are available upon reasonable request to the corresponding author

## Acknowledgments

The authors would like to thank the support of Universidade Federal of Paraná and the Postgraduate Program in Internal Medicine and Health Sciences.

## Data availability statement

Raw data are available upon reasonable request to the corresponding author.

## Appendices

The details of the model of the first-stage analysis are explained as follows:

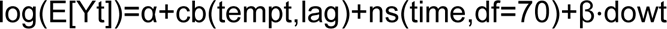

Where:

- E[Yt] represents the expected hospitalization count on day t.
- α is the model intercept.
- cb(tempt, lag) is the cross-basis matrix generated by the crossbasis function in the DLNM framework, which simultaneously models the exposure-response and lag-response relationships. The specification for this matrix was:

- Exposure-Response Relationship (Temperature): A quadratic B-spline (degree=2) with 3 internal knots (nk=3) placed at equally spaced values across each city’s temperature range.
- Lag-Response Relationship (Lag): A natural cubic B-spline for the cumulative lag effect, with a maximum lag of 7 days (lag=7) and 3 internal knots (nk=3) placed on a logarithmic scale to provide more flexibility to the initial lag days.
- ns(time,df=70) is a natural cubic spline with 70 degrees of freedom (equivalent to 10 per year, a standard approach for robust trend control) for the time variable, adjusting for long-term trends and seasonality.
- β⋅dowt represents the adjustment for the day-of-the-week effect (dow), included as a categorical variable.

